# Innate Immune Response Modulation and Resistance to SARS-CoV-2 infection: A Prospective Comparative Cohort Study in High Risk Healthcare Workers

**DOI:** 10.1101/2020.10.20.20214965

**Authors:** Sarita Rani Jaiswal, Anupama Mehta, Gitali Bhagwati, Rohit Lakhchaura, Hemamalini Aiyer, Bakulesh Khamar, Suparno Chakrabarti

## Abstract

To evaluate ability of modulated innate immune response to provide resistance to development of symptomatic RT-PCR confirmed COVID-19, 96 inpatient front line health care workers (HCW) were cohorted in 1:2 ratio to receive TLR2 agonist (heat killed Mycobacterium w, Mw; n=32) as innate immune response modulator or observation (n=64). All were followed up for 100 days. The incidence of COVID-19 was 31 (32.3%) for the entire cohort, with only one developing COVID-19 in Mw group (3.1% vs 46.8%. protective efficacy - 93.33%, p=0.0001; 95% CI 53.3-99.1). Self-limiting local injection site reaction was the only side effect and was seen in 14 HCW. Findings from the study suggest the potential for providing resistance against novel pathogen like SARS-CoV-2 by modulating innate immune response.

## Introduction

Front-line healthcare workers (HCW) are at the highest risk of acquiring COVID-19 with a 30-day hazard ratio (HR) of 24.3, compared to the general community^1^. Innate immune response is a conserved, prompt mechanism, which resists infections by recognizing the conserved pathogen-associated molecular patterns (PAMP) of an infectious agent by pathogen recognizing receptors (PRR) like toll like receptors (TLR). It is relevant in infections by novel pathogens like SARS-COV-2^2^. A TLR2 agonist, heat killed Mycobacterium w (Mw), also known as Mycobacterium Indicus Pranii, is, an approved immunomodulator in India^3^ and is used in the management of leprosy and other conditions. In a prospective open label cohort control study, we explored the efficacy of Mw in inducing resistance to the development of COVID-19 infection in HCW at a high risk of exposure, in a tertiary-healthcare set-up.

## Methods

Thirty-two HCWs from the Department of Blood and Marrow Transplantation and Hematology were administered 0.1 ml Mw (Sepsivac, Cadila Pharmaceuticals, India) intradermally in each arm on day 1 of the study (Mw group) and followed up for 100 days. 64 age matched HCWs from the rest of the hospital were enrolled in a control group. All HCW included in the study had a nasopharyngeal swab evaluated for SARS-CoV-2 by reverse transcriptase-polymerase chain reaction (RT-PCR), on development of symptoms suggestive of COVID-19 or following exposure to an infected person. Body temperature, pulse rate, oxygen saturation and self-reporting of symptoms was evaluated before and after each working day in the Mw group. ‘Exposure’ was defined as close contact with SARS-CoV-2 infected individuals without full protective gear. COVID-19 was graded as mild, moderate or severe as per WHO criteria. Subjects in the Mw group underwent two additional random SARS-CoV-2 specific RT-PCR evaluation 4 weeks apart. The study was approved by the institutional ethics committee.

## Results

The characteristics and outcomes of the Mw and control groups are detailed in Table 1. Overall, 31 out of 96 HCW enrolled had RT-PCR-confirmed COVID-19 infection of which 30 (96.77%) were in control group. Of the 31, who developed COVID-19 infection, four required hospitalization. All belonged to the control group. Despite a greater number of exposures in the Mw group, only one out of 32 (3.13%) subjects had an RT-PCR confirmed mild COVID-19 infection. HR for developing COVID-19 in the control group compared to the Mw group was 19.025 (p=0.0038). Based on this study, the resistance to infection (protective efficacy) provided by Mw was 93.33% (p=0.0001; 95% CI 53.3-99.1).

**Table 1:** Characteristics and outcome of study and control subjects in Mw Prophylaxis study.

|  | **Study Group (N=32)** | **Control Group (N=64)** | **P Value** |
| --- | --- | --- | --- |
| Age, Median, (Range), Years | 28 (22-56) | 28 (23-55) | 0.7 |
| Female/Male | 23/9 | 35/29 | 0.36 |
| No of Exposure, Median, (Range) | 2 (2-10) | 0 (0-2) | 0.0001 |
| No of RT-PCR Tests | 2 (2-4) | 1 (0-2) | 0.019 |
| RT-PCR Positive | 1 | 30 | 0.0001 |
| COVID-19<br>Mild/Mod/Severe | 1/0/0 | 26/4/0 | 0.0001 |
| Hospitalization | 0 | 4 | 0.36 |
| Duration of Symptoms,<br>Median (Range), Days | 3 | 12 (3-36) | 0.0001 |
| Mortality | 0 | 0 | 1.0 |

The only side effect noted with Mw was injection site reactions (moderate to severe - 4; mild - 10), which were self-limiting and did not require any specific management.

## Discussion

The study period coincided with the peak of the pandemic in New Delhi and the high rate of COVID-19 seen in control group is in line with that reported in similar HCW^1^. The resistance to COVID-19 seen in the Mw group suggests that a TLR2 agonist Mw might be useful in providing protection to subjects at a high risk of exposure. It will be interesting to study its long-term protective efficacy. BCG, another approved immunomodulator is also being evaluated for the prevention of COVID-19, and it will be interesting to study its outcome as there are differences in the innate immune response generated by the two in terms of being a TLR agonist and in ligand presentation^4^. Specific immune changes like upregulation of adaptive natural killer cells are being investigated by our group^5^ to understand the immune mechanism responsible for resistance to COVID-19. This study provides an initial proof to the concept of modulating the innate immune response for providing resistance to novel pathogens like SARS-COV2 until the availability of vaccines.

## Data Availability

Available on request.

